# Intracranial arteriosclerosis and cerebrovascular function in the general aging population – A 7T MRI Study

**DOI:** 10.1101/2025.05.21.25327070

**Authors:** A.M. Streiber, N.A.C. Kuenen, S.D.T. Pham, J. Neitzel, D. Bos, J.C.W. Siero, J.J.M. Zwanenburg, N. Dieleman, G.J. Biessels, M.W. Vernooij

## Abstract

**Background and Objectives:** Intracranial arteriosclerosis is a prevalent condition linked to serious health outcomes, such as stroke and dementia. It is hypothesized that intracranial arteriosclerosis compromises brain health by disrupting the functioning of adjacent vessels, including the middle cerebral artery (MCA) and the perforating small arteries downstream. Using 7T MRI, this study aims to assess the association between intracranial arteriosclerosis and cerebrovascular function in the MCA and in the downstream small perforating arteries.

**Methods:** This study is based on 195 participants (43.6% female, average age 70.8 years (± 4.52 years)) from the population-based Rotterdam Study who previously underwent a non-enhanced low-dose CT on which we measured intracranial carotid artery calcification (ICAC) and vertebral artery calcification (VAC) bilaterally as hallmark of intracranial arteriosclerosis. Participants underwent a 7T brain MRI to assess blood flow velocity and pulsatility in the MCA and small perforating arteries as well as whole-brain cerebrovascular reactivity (CVR). We assessed the relationship of left and right intracranial arteriosclerosis presence, burden, and subtypes with vessel function using linear regression and linear mixed models with a random intercept. Analyses were adjusted for age, sex and cardiovascular risk factors.

**Results:** The prevalence of ICAC was 77.4%. VAC prevalence was notably lower, at 10.3%. ICAC and VAC presence and burden were associated with higher pulsatility but not blood flow velocity in the MCA (e.g. β_ICAC presence_: 0.07 [95% Confidence interval: 0.02, 0.13]). There was no association with pulsatility or blood flow velocity in the small perforating arteries. Neither ICAC nor VAC were associated with CVR.

**Conclusions:** Intracranial arteriosclerosis influences pulsatility but not blood flow velocity in the MCA. This finding, combined with the absence of an effect between intracranial arteriosclerosis and cerebrovascular function further downstream, indicates that its impact is limited to cerebrovascular hemodynamics in adjacent vessels.

## INTRODUCTION

Intracranial arteriosclerosis is a highly prevalent pathology associated with serious health consequences including stroke and dementia [1-3]. However, the exact contribution of intracranial arteriosclerosis in disease development remains incompletely understood, limiting effective prevention and treatment strategies.

The pathways linking arteriosclerosis to these adverse clinical outcomes are multifaceted, involving factors such as small vessel disease [3] and inflammation [4, 5]. Recent studies have shown that intracranial arteriosclerosis is associated with accelerated cognitive decline [6] and imaging markers of vascular brain health, such as enlarged perivascular spaces, white matter hyperintensities, or lacunes [7, 8]. However, these markers typically reflect an end-stage and provide limited insight into the early functional changes in the adjacent arteries and the more distal, small vessels. It has been hypothesized that arteriosclerosis may impact the function of both the middle cerebral artery (MCA) and small vessels downstream [9]. These changes in vascular function may potentially contribute to small vessel disease or blood-brain barrier leakage, increasing the risk for dementia, stroke and mortality [10, 11]. Yet, the extent to which this hypothesis holds remains uncertain.

Vascular function is a broad term that encompasses several key measures, including pulsatility, blood flow velocity, and cerebrovascular reactivity (CVR). Pulsatility refers to the variability of blood flow velocity within vessels across the cardiac cycle and has been associated with small vessel disease, arteriosclerosis, and cognition [12-14]. Blood flow velocity describes the speed at which blood moves through the vessels and may be reduced in individuals with arteriosclerosis or a high cardiovascular disease risk profile, possibly due to widening of the arteries [15, 16]. CVR, a dynamic measure of vascular function, reflects the ability of blood vessels to dilate or constrict in response to vasoactive stimuli such as changes in blood CO_2_ levels [17]. Reduced CVR has been linked to modifiable risk factors for dementia and is thought to play a role in cerebrovascular pathology [18, 19].

Collectively, these vascular measures provide insight into the health and functionality of the cerebral vasculature and have been previously linked to stroke and cognitive decline [20-22]. However, the role of intracranial arteriosclerosis in these downstream cerebrovascular processes remains largely unexplored. Only one previous pilot study investigated this association in symptomatic cerebral small vessel disease patients with a history of lacunar infarcts and deep intracerebral hemorrhage [23]. Our goal is to assess this relationship further in a larger, population-based study with a wider variability of vascular pathology.

Due to the technical properties of commonly used magnetic resonance imaging (MRI) scanners, measuring function in the cerebral microvasculature is challenging [21]. However, high field strength imaging with 7T MRI now facilitates the direct high-resolution *in vivo* assessment of structural and functional neurovascular markers, providing further understanding of microvascular changes in early stages [24]. Using 7T MRI, we aim to comprehensively assess the association between intracranial arteriosclerosis and cerebrovascular functioning, first by examining vessel function of the MCA and, then by extending this investigation to small perforating arteries further downstream.

Firstly, we hypothesize that the presence and burden of intracranial artery calcification, as a hallmark of intracranial arteriosclerosis, is associated with higher pulsatility and lower mean blood-flow velocity of the MCA and of small perforating arteries in the basal ganglia and semioval center. Additionally, we hypothesize that intracranial arteriosclerosis is associated with lower whole-brain CVR.

Secondly, we propose that arteriosclerosis on one hemispheric side has a stronger association with small vessel function on the ipsilateral side than the contralateral side, as small vessels on the same side are more directly affected by the downstream impact of the pathology.

In an additional exploratory analysis, we will investigate if subtypes of intracranial arteriosclerosis differentially affect downstream vascular function.

## METHODS

### Study Participants

The current study is part of the population-based Rotterdam study, which was implemented in 1990 to investigate the prevalence and determinants of common age-related disease in Rotterdam, the Netherlands [25]. All Rotterdam Study participants receive one extensive baseline examination followed by regular study visits every three to six years. The current study sample consists of 195 stroke- and dementia-free study participants who underwent a 7T brain MRI to assess small vessel function. As part of a previous substudy investigating the interplay between amyloid- and vascular pathology, all participants underwent an amyloid positron emission tomography–computed tomography (PET-CT) scan two to three years before the 7T brain MRI. The detailed in- and exclusion criteria have been mentioned in greater detail elsewhere [26]. A detailed overview of the participant selection is provided in Figure 1.

**Figure 1.**
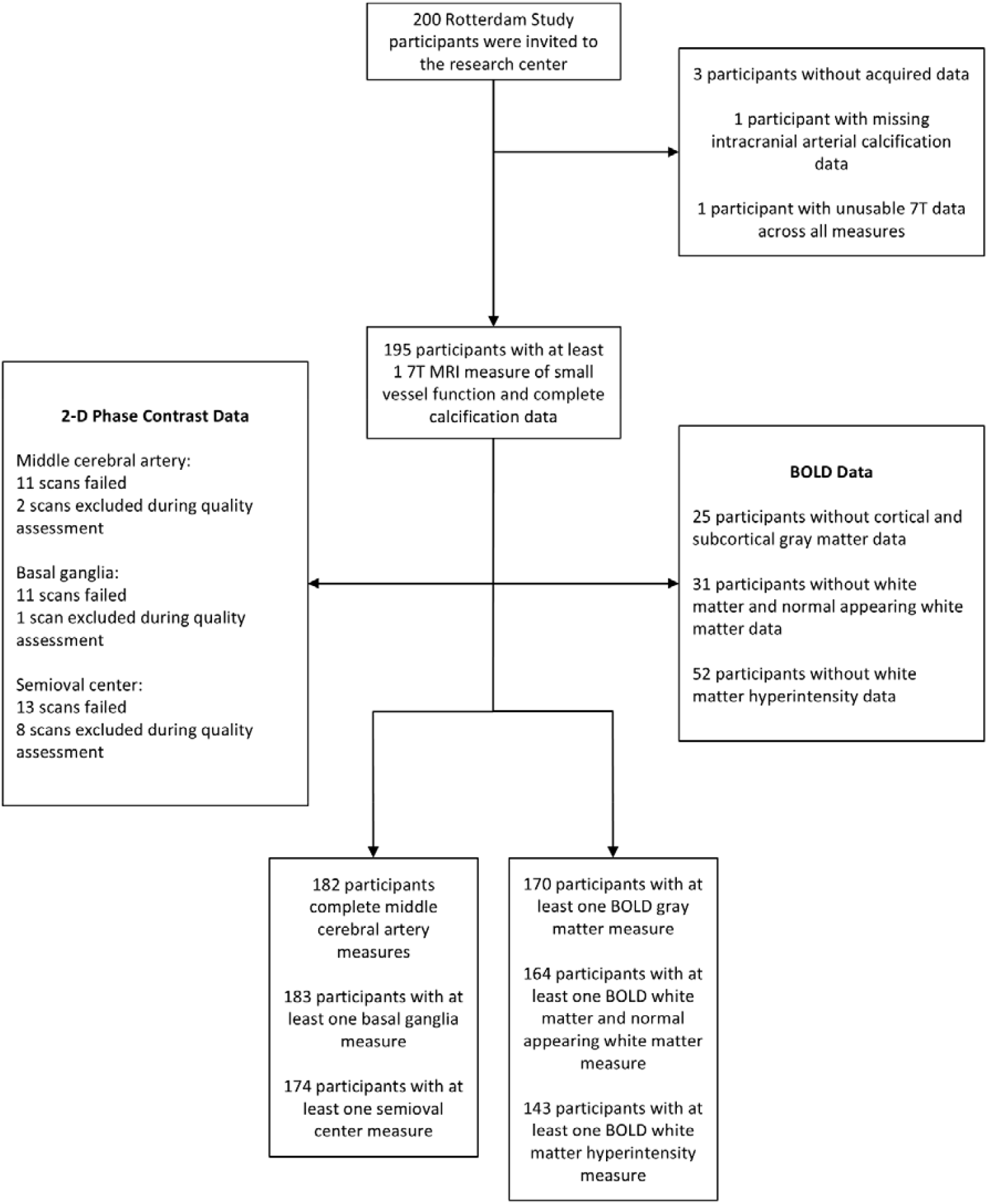
Flowchart of participant selection. Participants with middle cerebral artery data were included only if they had a complete measurement, as this parameter was assessed unilaterally. Since all other measures were bilateral, having at least one usable measurement was sufficient to be included in the analysis.

### Arteriosclerosis Assessment

Using the anatomical low-dose CT scan from the amyloid PET-CT, we measured intracranial carotid artery calcification (ICAC) and vertebral artery calcification (VAC) bilaterally. These measures served as markers for intracranial arteriosclerosis. The exact scoring procedure has been described elsewhere [27]. In brief, two trained independent raters manually quantified ICAC and VAC in the anterior and posterior circulation. ICAC was evaluated bilaterally from the petrous intracranial carotid artery to the sella turcica, while VAC was assessed in the vertebral arteries. Calcification volumes (in mm^3^) were calculated using a validated semiautomatic scoring method [28], based on regions of interest drawn around calcifications and applying a threshold of 130 Hounsfield Units.

ICAC subtypes were categorized into two groups based on their circularity, thickness, and morphology: predominantly intimal calcification, and predominantly internal elastic lamina (IEL) calcification. These subtypes are based on previous research [29]. Since standardized tools for VAC subtypes have not yet been validated and VAC prevalence is generally low [30], only ICAC subtypes were included in the final analysis.

### Blood Flow Velocity and Pulsatility Assessment

All cerebrovascular artery measures have been obtained using a Philips 7T scanner (Philips Healthcare, Best, The Netherlands) with a 32-channel receive head coil in combination with a quadrature transmit coil (Nova Medical, MA, USA). The detailed scan parameters and protocols can be found elsewhere [31]. The mean time difference between the calcification assessment and the 7T scan was 2.7 years (SD=0.7 years).

Cerebral artery flow data of the MCA and of the small perforating arteries at the level of the basal ganglia and the semioval center, were acquired using three single-slice 2D phase-contrast acquisitions. MCA flow data were analyzed using Small vessEL MArker (SELMA) analysis software, an open-source tool for artery flow velocity analyses [32]. First, the researchers manually delineated regions of interest on the single 2D slice planned perpendicularly on the MCA (supplemental Figure S1). Subsequently, voxel-wise noise estimation was performed, followed by the computation of magnitude and velocity signal-to-noise ratios. Arteries were identified based on signal-to-noise ratio thresholds, with automated filtering applied to exclude vessels that were not perpendicular to the scanning plane and to remove ghosting artifacts. To ensure that only the MCA was included in this slice, only the largest artery with the highest velocity was selected and all other detected arteries, most likely smaller perforating arteries or branches of the MCA, were discarded.

Perforating artery flow downstream was assessed using two slices positioned at the basal ganglia and semioval center based on anatomical landmarks. Apparent vessel detections in ghosting artefacts in the semioval center and perforating arteries in the basal ganglia that were oriented non-perpendicularly to the scanning plane have been excluded automatically using a previously developed method [31]. Additionally, apparent perforating arteries that were located within a 1.2 mm radius from each other have been excluded, as these mostly are ‘false detections’ of larger and non-perpendicular vessels.

Mean blood flow velocity of the MCA and the small perforating arteries was quantified as the averaged mean velocity per subject in cm/s. Subsequently, the blood flow velocity pulsatility index (PI) was calculated individually as 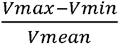 with V_max_, V_min_, and V_mean_ indicating the maximum, minimum and mean of the normalized and averaged blood flow velocity, respectively. Blood flow data were acquired unilaterally for the MCA. The side for MCA data acquisition was based on which side provided the best planning perpendicular to the vessel orientation. In case both sides were equally well aligned, one side was randomly selected. Blood flow data for the small perforating arteries were acquired bilaterally.

### Cerebrovascular Reactivity Assessment

Whole-brain CVR was measured using blood-oxygenation level-dependent contrast-weighted MRI during exposure to a hypercapnic stimulus [31]. Participants wore a face mask through which they breathed medical room air and 6% CO_2_ in air with a flow rate of 30 L/min in a gas challenge paradigm alternating two two-minute blocks of CO_2_ with three two-minute blocks of medical room air. Gasses have been pre-mixed in medically graded cylinders (Linde Gas, The Netherlands). Monitoring equipment recorded pulse rate (sampling rate = 500 Hz) and end-tidal CO_2_ (etCO2, 40 Hz; CD3-A AEI Technologies, Pittsburgh, USA). The blood-oxygenation level-dependent CVR was computed using a general linear model with the end-tidal CO_2_ used as regressor. In line with earlier studies [33], CVR was normalized with the ΔCO_2_ in mmHg obtained from the end-tidal CO_2_ traces. CVR amplitudes (in %/mmHg) have been assessed in the cortical and subcortical gray matter, and total white matter, normal-appearing white matter and white matter hyperintensities.

### Assessment of Covariables

Besides intracranial artery calcification, pulsatility and blood flow velocity, we measured age, sex, body mass index, diabetes, smoking status (never, former, current), hypertension, and dyslipidemia. The body mass index was computed as weight (in kg) divided by height^2^ (in m). Diabetes was defined as fasting serum glucose levels of ≥ 7mmol/L and/or use of antidiabetic medication. Participants were rated as hypertensive if their systolic blood pressure was ≥140 mmHg, if their diastolic blood pressure was ≥90 mmHg, and/or if they used antihypertensive medication. Dyslipidemia was defined as having a total cholesterol concentration of at least 6.2 mmol/L and/or using lipid-lowering medication. All variables were measured at the most recent Rotterdam Study visit approximately nine months before the 7T MRI scan.

### Statistical Analyses

We first examined the descriptive characteristics of the study sample and summarized all determinants, outcomes, and covariates. To provide a comprehensive overview of pulsatility and blood flow velocity in the MCA and the small perforating arteries further downstream, we additionally computed Spearman correlation coefficients. To examine the relationship between intracranial arteriosclerosis and pulsatility and blood flow velocity in the MCA, we applied multiple linear regression models. To assess the relationship between intracranial arteriosclerosis and pulsatility, blood flow velocity, and CVR in the small perforating arteries, we used linear mixed models with a random intercept to account for within-subject correlation between the two-sided measurements. One participant had missing smoking data at the time of the study visit, which was imputed using the last observation carried forward method. We imputed this value using the most recent available observation of a previous study visit, which was collected in 2007.

The ICAC burden has been quantified by temporarily excluding participants without ICAC from the dataset and subsequently assigning calcification tertiles to all participants with prevalent ICAC. Following the tertile computation, participants without ICAC were added back to the dataset and received a score of 0. The resulting ICAC variable was dummy coded to compare participants in tertiles 1, 2, and 3 to participants without calcification. Similarly to ICAC tertiles, ICAC subtypes were dummy coded and compared to participants without calcification. In contrast to ICAC, the VAC burden has been quantified using the raw calcification volume (in mm^3^). Due to its low prevalence, the tertile computation would have led to small subgroups, limiting statistical power.

Determinants of middle cerebral artery- and small vessel functional markers were calcification presence (yes/ no), calcification burden (in tertiles for ICAC and in mm^3^ for VAC), and ICAC subtypes. For MCA analyses, the ICAC subtype categorization for the left and right sides was adapted because MCA pulsatility was measured on only one side. Participants were classified as having predominantly intimal or IEL calcification if the respective subtype was present bilaterally. A mixed category was introduced for cases with intimal calcification on one side and IEL calcification on the other. For the small vessel analyses using linear mixed models, only two subtype categories (intimal and IEL) were used, as they were combined with bilateral outcome measures. In all analyses, subtypes were compared to the absence of calcification.

The main outcomes of this study included blood flow velocity in cm/s and pulsatility indices in the MCA, basal ganglia, and semioval center, as well as CVR in cortical and subcortical gray matter, total white matter, normal-appearing white matter and white matter hyperintensities. The CVR distribution for white matter hyperintensities was strongly skewed and log-transformed to fulfil the normality and homoscedasticity assumptions underlying the regression models. We first adjusted all regression models for age and sex (model 1). In a next step, all models were additionally adjusted for the time difference between PET-CT and 7T MRI, body mass index, diabetes, smoking status, hypertension, and dyslipidemia (model 2). Regression estimates reflect the change in small vessel function markers based on calcification presence and burden.

To assess correlations between intracranial arteriosclerosis, pulsatility, blood flow velocity, and cerebrovascular reactivity separately for the left and right hemisphere, we computed Spearman correlation coefficients between all lateralized determinants and outcomes. These correlation analyses could not be carried out for MCA measures as they were only obtained unilaterally.

To investigate the potential impact of selection bias on our results, we compared all participant characteristics to those from another Rotterdam Study cohort with available CT data but who did not undergo 7T MRI [34]. All analyses have been carried out in R version 4.2.2 [35].

### Data Availability

This study follows the STROBE (Strengthening the Reporting of Observational Studies in Epidemiology) reporting guidelines [36]. Data can be obtained upon request. Requests should be directed towards the management team of the Rotterdam Study, which has a protocol for approving data requests. Because of restrictions based on privacy regulations and informed consent of the participants, data cannot be made freely available in a public repository. The Rotterdam Study has been approved by the Medical Ethics Committee of the Erasmus MC (registration number MEC 02.1015) and by the Dutch Ministry of Health, Welfare and Sport (Population Screening Act WBO, license number 1071272-159521-PG). The Rotterdam Study has been entered into the Netherlands National Trial Register (NTR; www.trialregister.nl) and into the WHO International Clinical Trials Registry Platform (ICTRP; https://apps.who.int/trialsearch/) under shared catalogue number NTR6831. All participants provided written informed consent to participate in the study and to have their information obtained from treating physicians.

## RESULTS

A descriptive overview of the study sample is presented in Table 1. The average age in this sample was 70.8 years (± 4.5 years) and 43.6% of the participants were female. The prevalence of ICAC was 77.4%, while the VAC presence was notably lower, at 10.3%. The correlation plot for pulsatility and blood flow velocity in the MCA and downstream small perforating arteries is summarized in supplemental Figure S2. Pulsatility in the MCA showed a weak correlation with pulsatility in the semioval center (r=0.23), with a stronger correlation observed in the basal ganglia (r=0.31). Similarly, blood flow velocity in the MCA was weakly correlated with blood flow velocity in the basal ganglia (r=0.16). We found no correlation between blood flow velocity in the MCA and the semioval center.

**Table 1.**
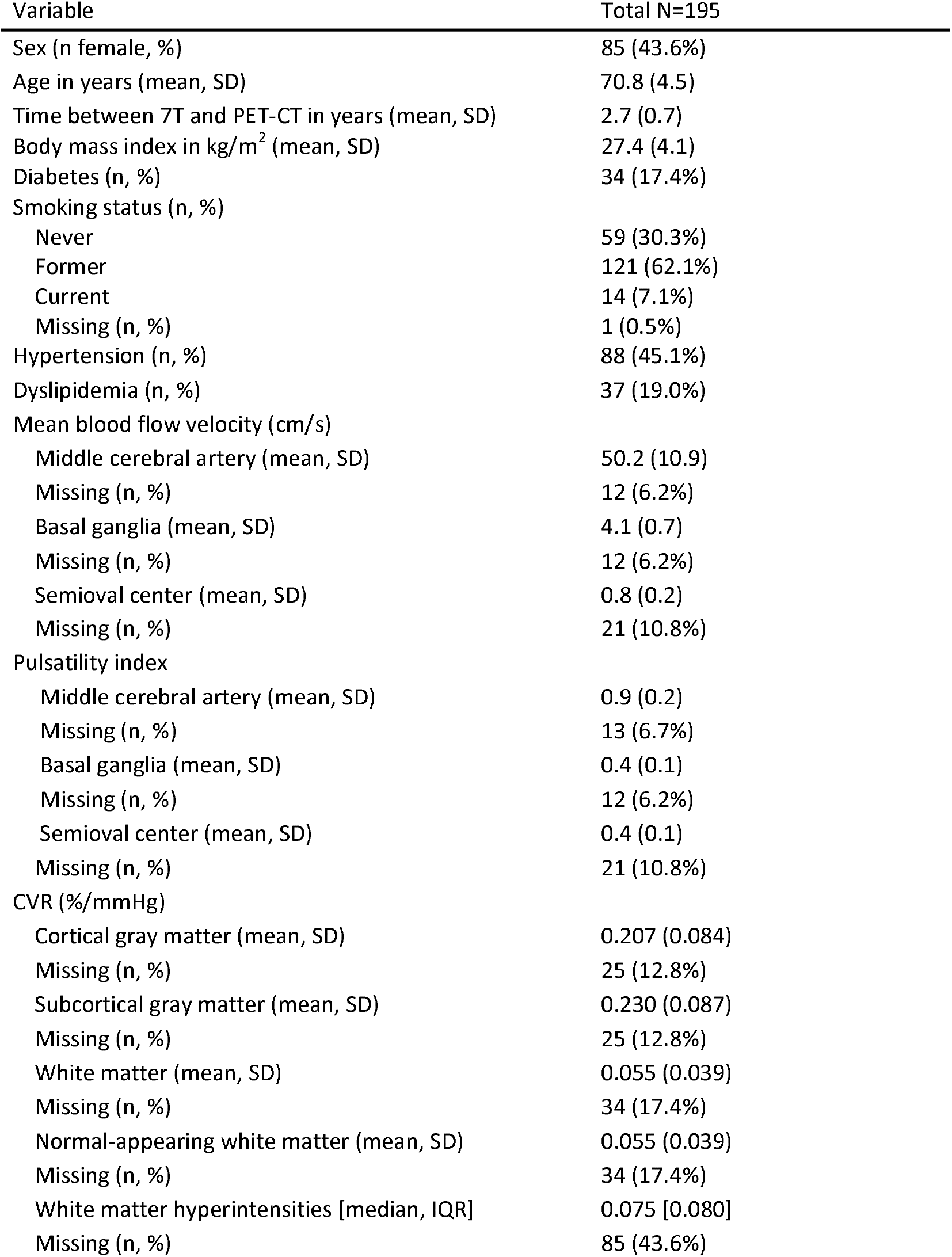
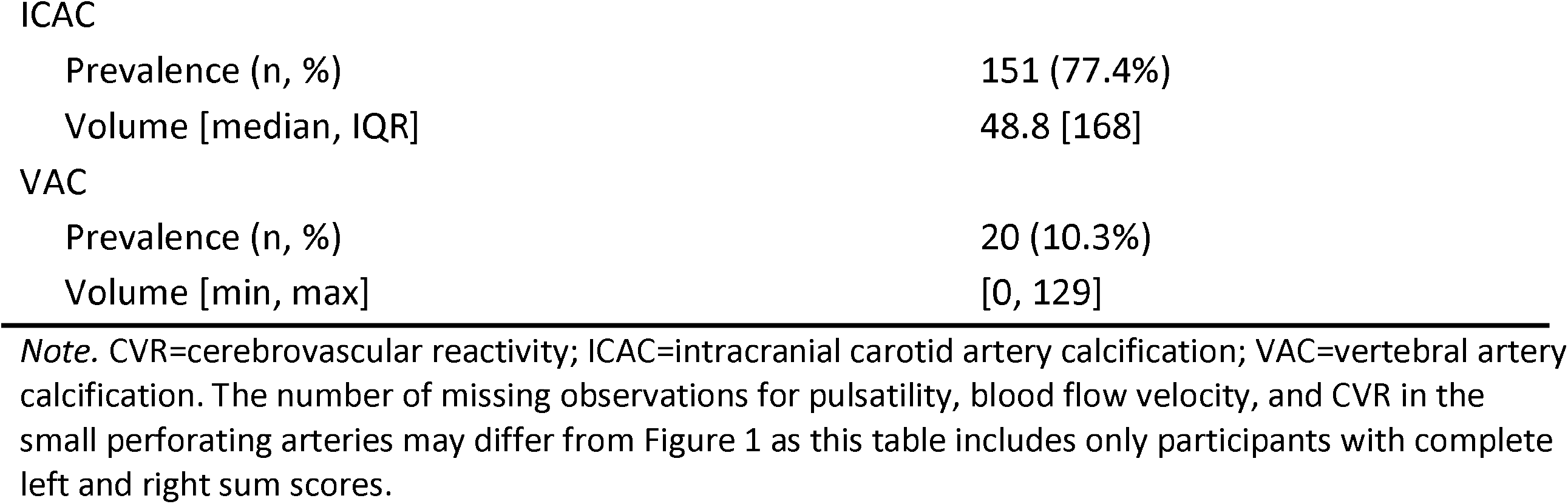
Demographic and clinical participant characteristics. Variable.

| Variable | Total N=195 |
| --- | --- |
| Sex (n female, %) | 85 (43.6%) |
| Age in years (mean, SD) | 70.8 (4.5) |
| Time between 7T and PET-CT in years (mean, SD) | 2.7 (0.7) |
| Body mass index in kg/m<sup>2</sup> (mean, SD) | 27.4 (4.1) |
| Diabetes (n, %) | 34 (17.4%) |
| Smoking status (n, %) |  |
| Never | 59 (30.3%) |
| Former | 121 (62.1%) |
| Current | 14 (7.1%) |
| Missing (n, %) | 1 (0.5%) |
| Hypertension (n, %) | 88 (45.1%) |
| Dyslipidemia (n, %) | 37 (19.0%) |
| Mean blood flow velocity (cm/s) |  |
| Middle cerebral artery (mean, SD) | 50.2 (10.9) |
| Missing (n, %) | 12 (6.2%) |
| Basal ganglia (mean, SD) | 4.1 (0.7) |
| Missing (n, %) | 12 (6.2%) |
| Semioval center (mean, SD) | 0.8 (0.2) |
| Missing (n, %) | 21 (10.8%) |
| Pulsatility index |  |
| Middle cerebral artery (mean, SD) | 0.9 (0.2) |
| Missing (n, %) | 13 (6.7%) |
| Basal ganglia (mean, SD) | 0.4 (0.1) |
| Missing (n, %) | 12 (6.2%) |
| Semioval center (mean, SD) | 0.4 (0.1) |
| Missing (n, %) | 21 (10.8%) |
| CVR (%/mmHg) |  |
| Cortical gray matter (mean, SD) | 0.207 (0.084) |
| Missing (n, %) | 25 (12.8%) |
| Subcortical gray matter (mean, SD) | 0.230 (0.087) |
| Missing (n, %) | 25 (12.8%) |
| White matter (mean, SD) | 0.055 (0.039) |
| Missing (n, %) | 34 (17.4%) |
| Normal-appearing white matter (mean, SD) | 0.055 (0.039) |
| Missing (n, %) | 34 (17.4%) |
| White matter hyperintensities [median, IQR] | 0.075 [0.080] |
| Missing (n, %) | 85 (43.6%) |
| ICAC |  |
| Prevalence (n, %) | 151 (77.4%) |
| Volume [median, IQR] | 48.8 [168] |
| VAC |  |
| Prevalence (n, %) | 20 (10.3%) |
| Volume [min, max] | [0, 129] |

*Note.* CVR=cerebrovascular reactivity; ICAC=intracranial carotid artery calcification; VAC=vertebral artery calcification. The number of missing observations for pulsatility, blood flow velocity, and CVR in the small perforating arteries may differ from Figure 1 as this table includes only participants with complete left and right sum scores.

The results of the fully adjusted multiple linear regression models for MCA pulsatility and blood flow velocity are summarized in Table 2. Model 1 adjusted for age and sex is presented in supplementary Table S1. Both presence and a higher burden of ICAC were associated with higher MCA pulsatility (β_Presence_: 0.07 [95% confidence interval (CI): 0.02, 0,13]; β_Tertile 2_: 0.09 [95% CI: 0.02, 0,15]; β_Tertile 3:_ 0.11 [95% CI: 0.04, 0,18]). Furthermore, both ICAC subtypes were associated with higher pulsatility. The point estimates were higher for IEL and mixed calcification (β_IEL_: 0.10 [95% CI: 0.03, 0.18]; β_Mixed_: 0.11 [95% CI: 0.03, 0.19]) than for intimal calcification (β_Intimal_: 0.06 [95% CI: -0.00, 0.11]), though the confidence intervals were largely overlapping. Similarly to ICAC, VAC presence was also associated with higher MCA pulsatility (β_Presence_: 0.08 [95% CI: -0.00, 0.15]). Neither ICAC nor VAC presence and burden were associated with blood flow velocity measures in the MCA.

**Table 2.** Linear regression results showing the relationship between intracranial arteriosclerosis and middle cerebral artery pulsatility and blood flow velocity.

| Coefficient | Pulsatility |  | Blood flow velocity |  |
| --- | --- | --- | --- | --- |
|  | Beta | 95% CI | Beta | 95% CI |
| ICAC presence | 0.07 | 0.02 - 0.13 | 1.14 | -2.48 - 4.76 |
| ICAC tertiles |  |  |  |  |
| 1 | 0.04 | -0.02 - 0.10 | 1.42 | -2.79 - 5.64 |
| 2 | 0.09 | 0.02 - 0.15 | -0.64 | -4.86 - 3.58 |
| 3 | 0.11 | 0.04 - 0.18 | 3.91 | -0.81 - 8.63 |
| ICAC subtypes |  |  |  |  |
| Intimal | 0.06 | -0.00 - 0.11 | 1.39 | -2.45 - 5.24 |
| IEL | 0.10 | 0.03 - 0.18 | -1.05 | -5.96 - 3.86 |
| Mixed | 0.11 | 0.03 - 0.19 | 3.18 | -2.16 - 8.52 |
| VAC presence | 0.08 | -0.00 - 0.15 | -2.16 | -7.22 - 2.91 |
| VAC volume | 0.00 | 0.00 - 0.00 | -0.05 | -0.16 - 0.07 |

*Note.* ICAC=intracranial carotid artery calcification; VAC=vertebral artery calcification;
IEL=internal elastic lamina calcification. Betas indicate the change in mean blood flow velocity (cm/s) for categorical calcification estimates compared to no calcification or per 1mm<sup>3</sup> increase of calcification. All models have been adjusted for age, sex, body mass index, diabetes, smoking status, hypertension, and dyslipidemia.

Table 3 and Table 4 show the results of the fully adjusted linear mixed models for pulsatility and blood flow velocity in the small perforating arteries, respectively. The complementary models adjusted for age and sex (Model 1) are presented in supplementary Table S2 and Table S3. Neither ICAC nor VAC measures were associated with pulsatility and blood flow velocity in the small perforating arteries. In Table 5, we summarized the fully adjusted linear mixed model results on the relationship between intracranial arteriosclerosis and whole-brain CVR. Results for Model 1 adjusted for age and sex are shown in supplementary Table S4. While none of the results were statistically significant, the point estimates indicate that both ICAC and VAC were consistently associated with lower CVR in the cortical and subcortical gray matter as well as in regions with white matter hyperintensities.

**Table 3.**
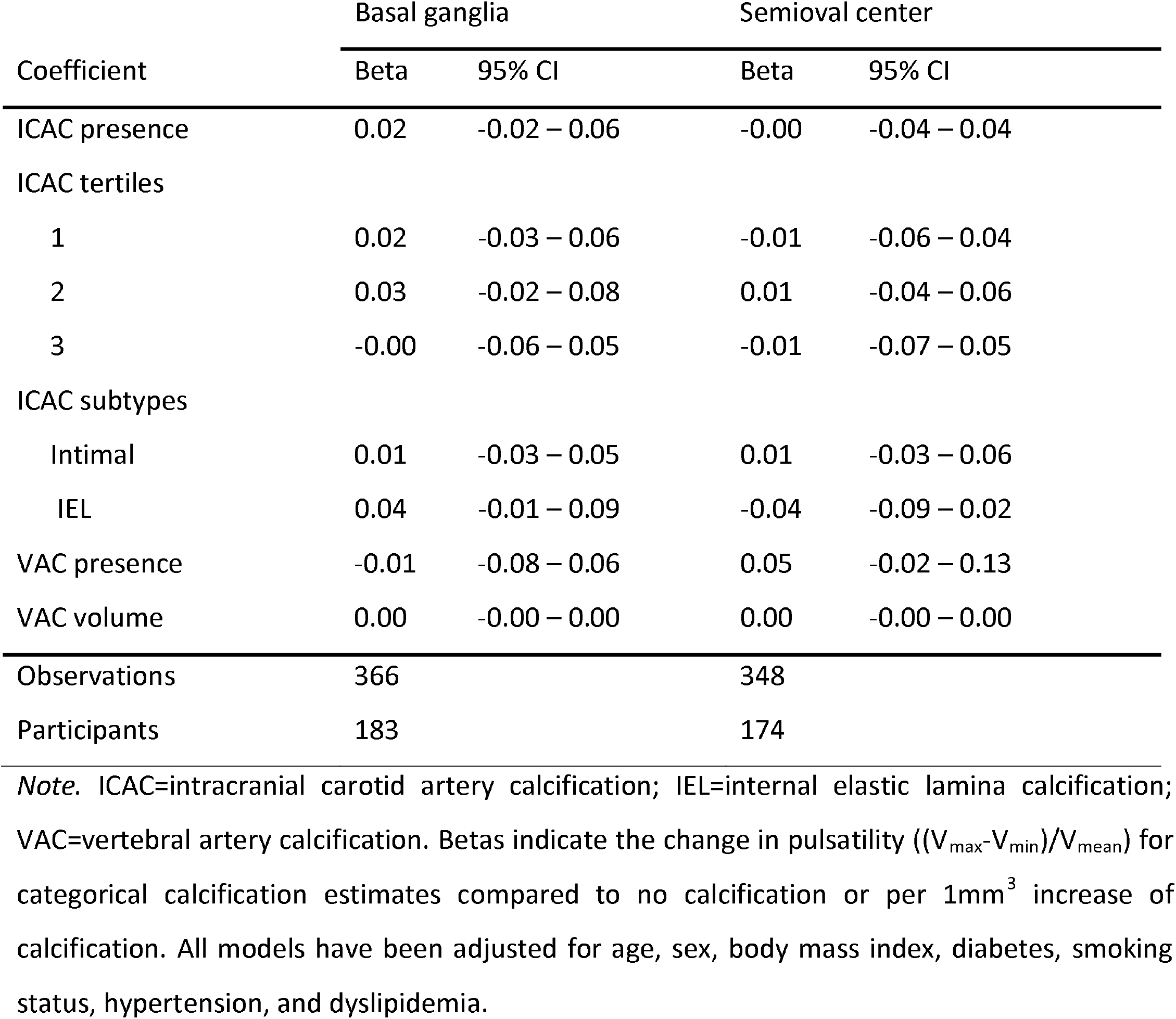
Linear mixed model results showing the relationship between intracranial arteriosclerosis and pulsatility in the basal ganglia and semioval center.

**Table 4.**
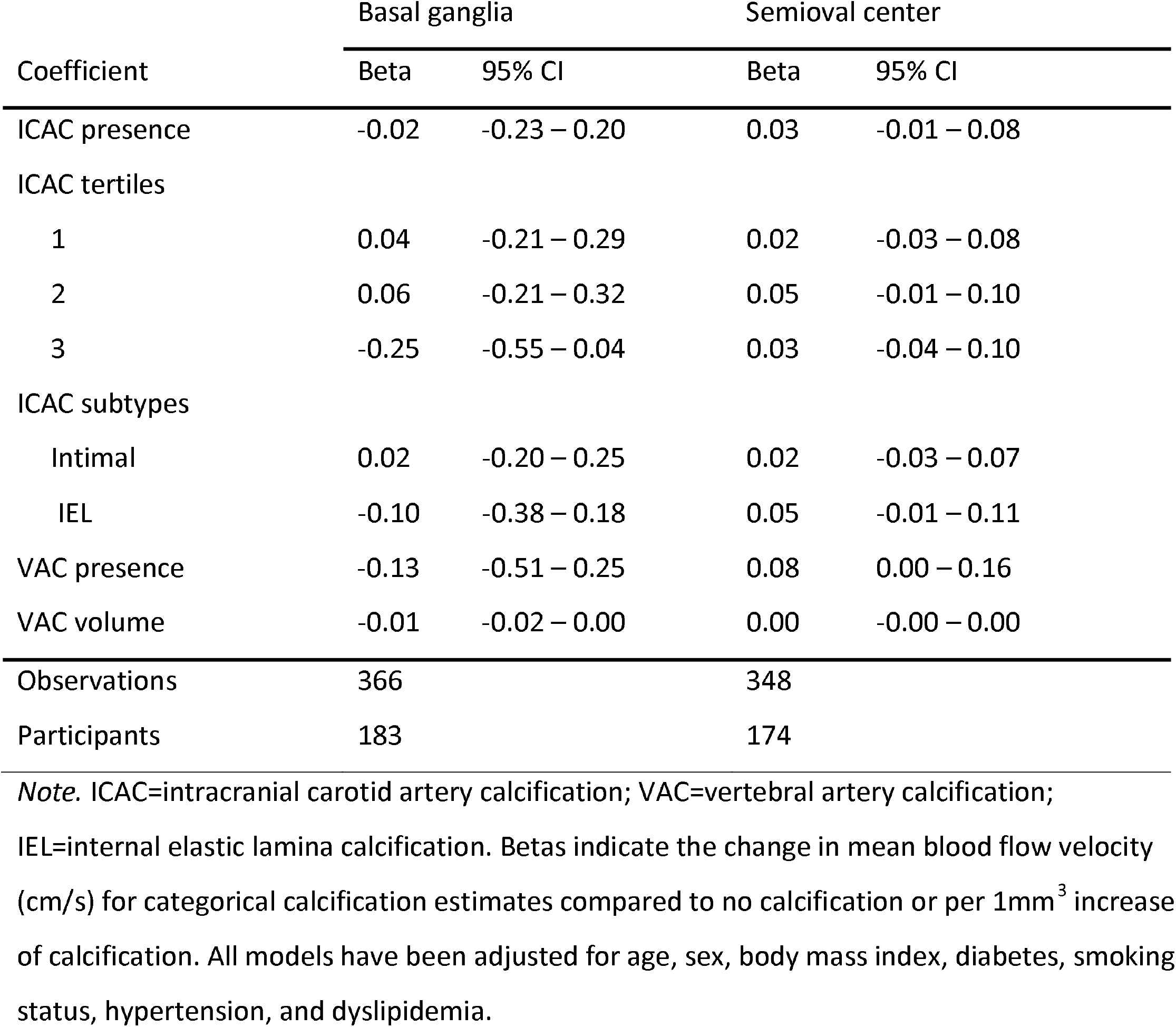
Linear mixed model results showing the relationship between intracranial arteriosclerosis and blood flow velocity in the basal ganglia and semioval center.

**Table 5.**
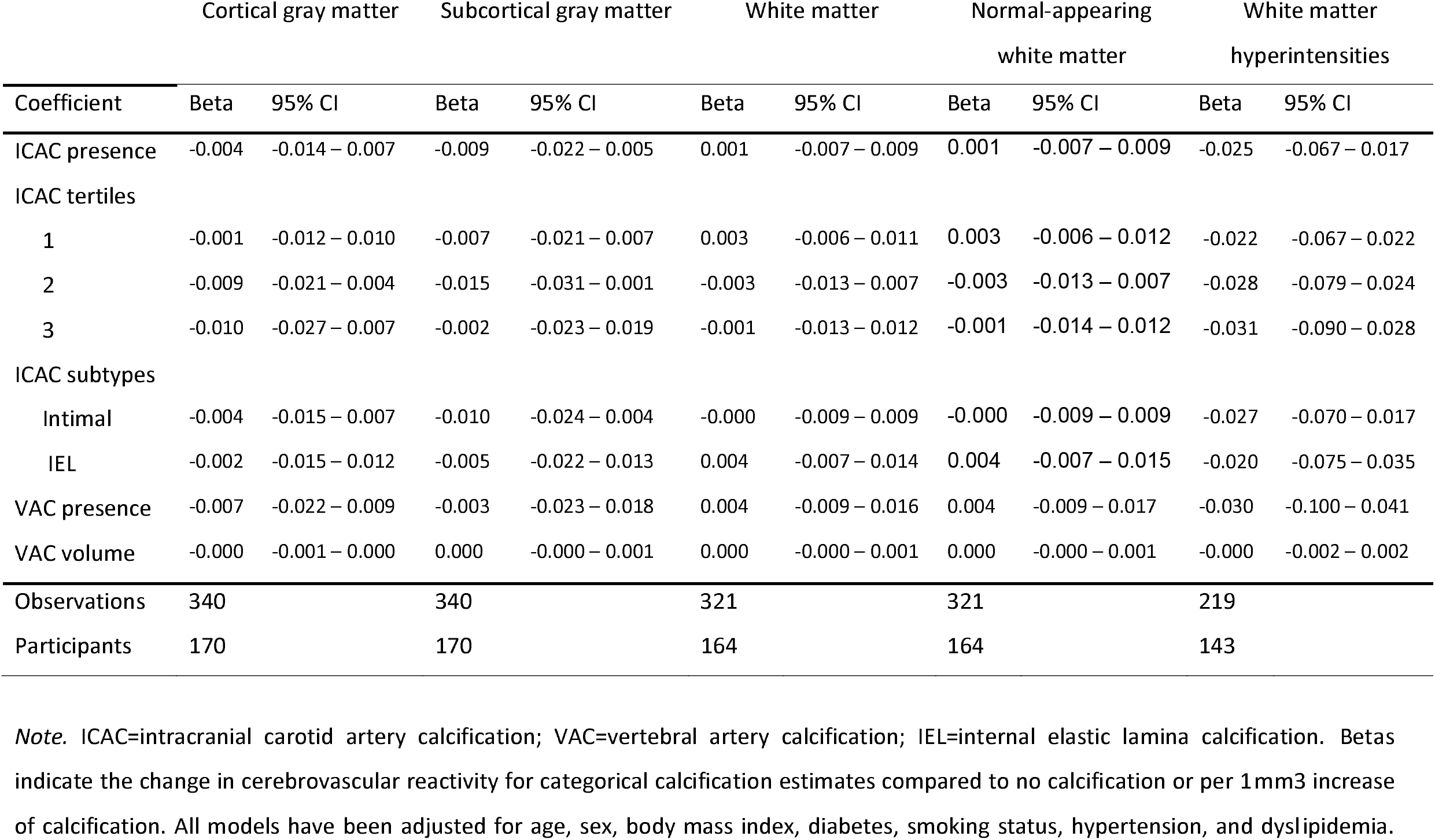
Linear mixed model results showing the relationship between intracranial arteriosclerosis and cerebrovascular reactivity.

The correlation coefficients indicated no differences between left and right ICAC and VAC regarding the correlation with ipsi- or contralateral small vessel function in the basal ganglia and the semioval center (Figure 2). Furthermore, the correlation analysis showed no differences between left and right arteriosclerosis estimates and their effect on CVR (Figure 3). Higher volumes of both left and right arteriosclerosis consistently, but non-significantly, correlated with a lower blood flow velocity, a higher pulsatility index, and a lower CVR. Overall, left and right estimates of the same modality were moderately to highly correlated.

**Figure 2.**
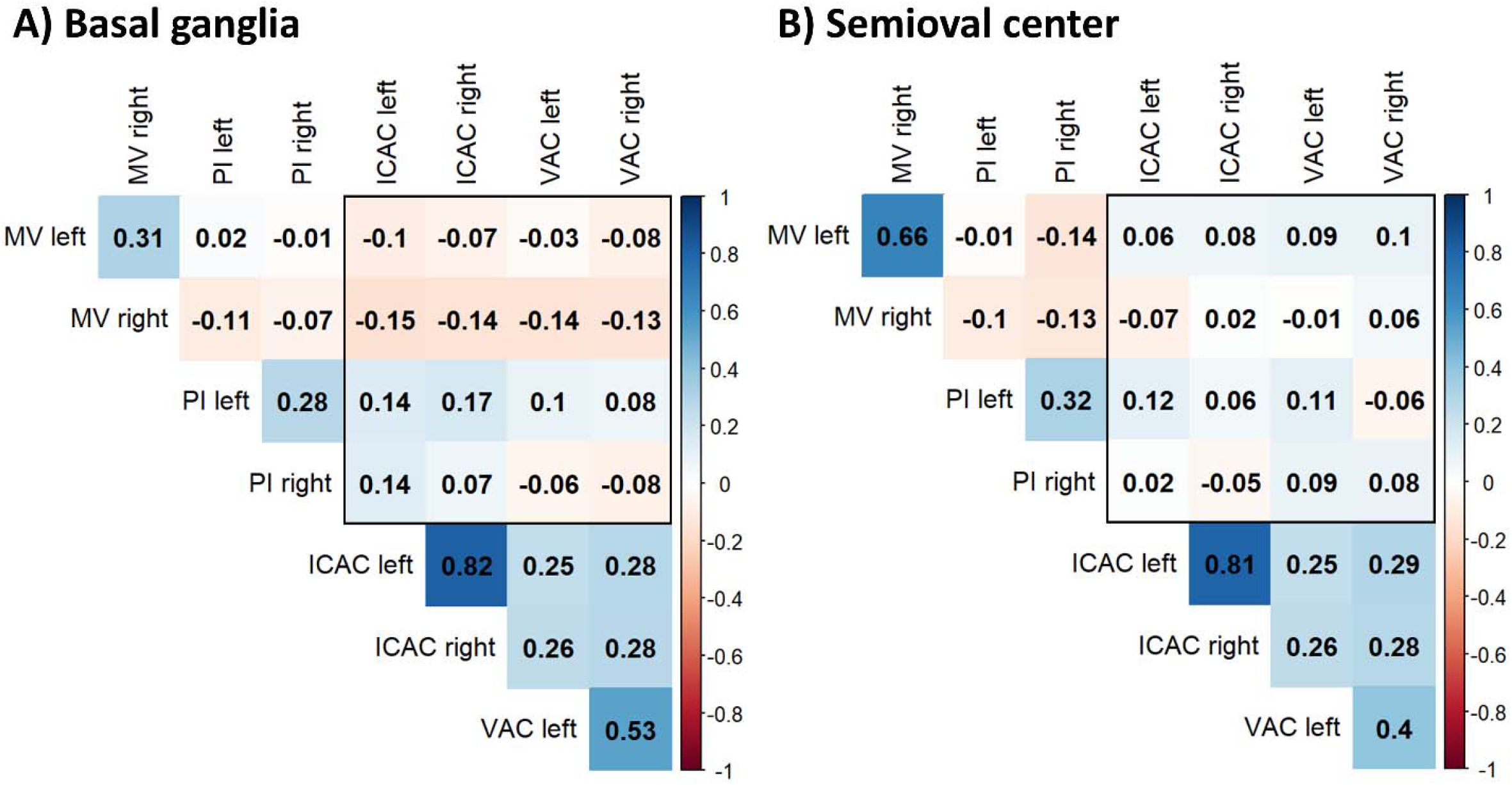
Unadjusted correlation coefficients for lateralized intracranial carotid artery calcification (ICAC), vertebral artery calcification (VAC), pulsatility (PI), and mean-blood flow velocity (MV) in the basal ganglia (A) and semioval center (B).

**Figure 3.**
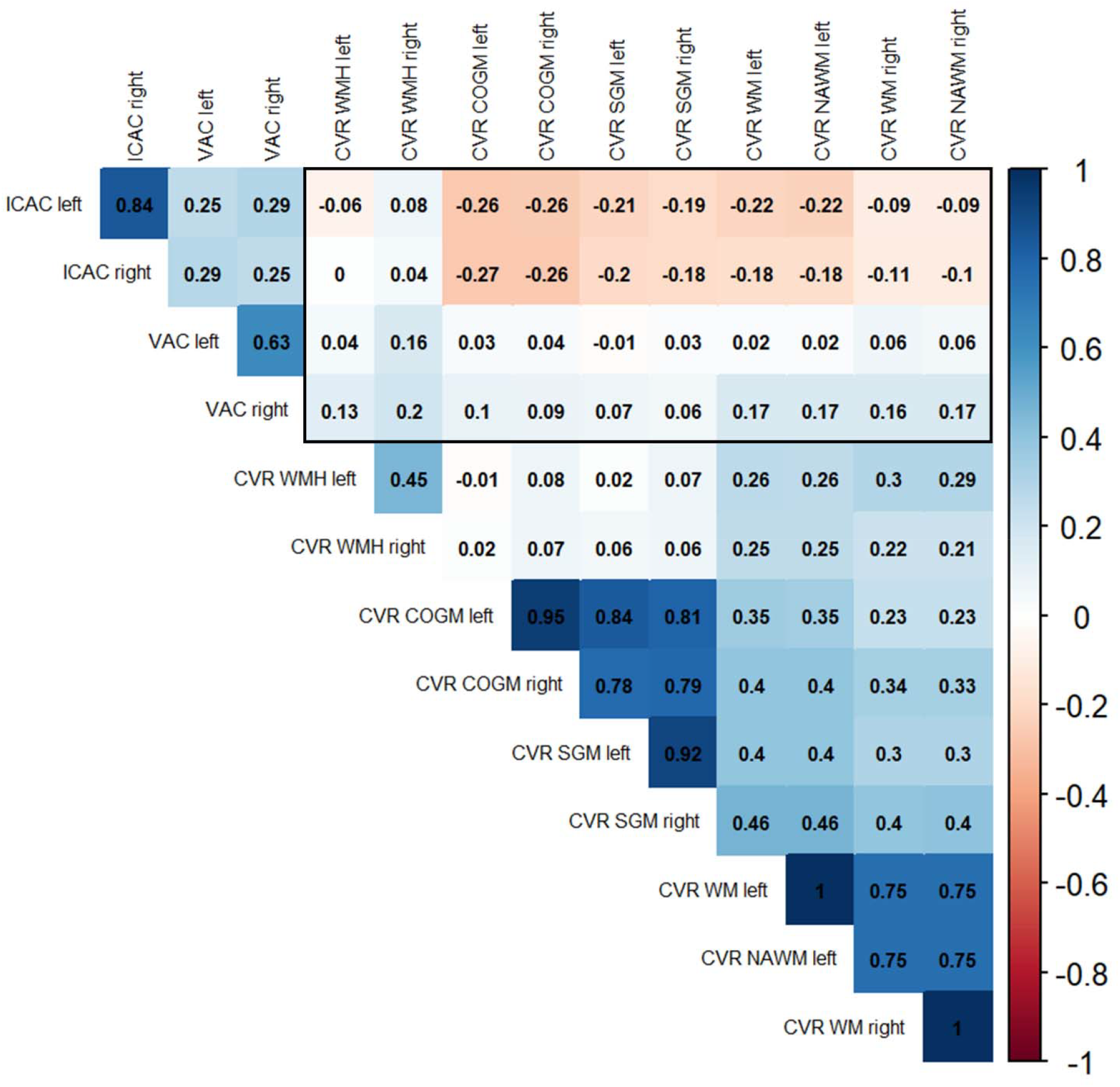
Unadjusted correlation coefficients for lateralized intracranial carotid artery calcification (ICAC), vertebral artery calcification (VAC), and cerebrovascular reactivity (CVR) in the cortical gray matter (COGM), subcortical gray matter (SGM), white matter (WM), normal-appearing white matter (NAWM), and white matter hyperintensities (WMHs).

### Sensitivity Analysis

To investigate potential selection bias, we compared descriptive characteristics of our current sample to those from an earlier Rotterdam Study cohort with available arterial calcification measures (Table S5). Compared to the age- and sex-matched sample from the Rotterdam Study, fewer participants in our study had hypertension (45.7% vs. 73.3%), dyslipidemia (19.0% vs 24.1%), or a history of smoking (69.8% vs. 72.8%). Additionally, the prevalence of ICAC and VAC was lower in our cohort relative to the other CT sample, with a VAC prevalence of 10.3% compared to 19.1%. There were no major differences regarding the mean BMI and the prevalence of diabetes between the two groups.

## DISCUSSION

In this study, we investigated the relationship between intracranial arteriosclerosis and cerebrovascular function in the middle cerebral artery and small perforating arteries using 7T MRI in the general aging population. Arteriosclerosis in the intracranial carotid arteries and vertebrobasilar arteries was consistently associated with higher pulsatility in the MCA, but not in the small perforating arteries. Overall, we did not observe a relationship of ICAC and VAC with blood flow velocity in the MCA and the perforating small vessels downstream. Furthermore, our analyses indicated that there was no statistically significant association between intracranial arteriosclerosis and whole-brain CVR. There was no indication that associations were stronger for ipsilateral arteriosclerosis compared to the contralateral side. The association between ICAC and MCA pulsatility is consistent with previous studies [9, 37], confirming that intracranial arteriosclerosis contributes to higher pulsatility in adjacent vessels. The relationship between VAC and MCA pulsatility, despite their anatomical distance, may be explained by their shared contribution to the Circle of Willis [38]. Additionally, vertebrobasilar artery calcification tends to occur in later stages of arteriosclerosis, often when individuals already exhibit significant ICAC [30]. This suggests that VAC may serve as a marker of disease severity, and the observed relationship between VAC and MCA pulsatility could reflect a more advanced stage of vascular pathology, which affects pulsatility downstream. Consequently, VAC may affect MCA pulsatility independently or in interplay with ICAC.

Compared to intimal calcification, both IEL calcification and mixed calcification patterns exhibited slightly higher point estimates for their association with MCA pulsatility. However, their corresponding 95% confidence intervals largely overlapped, indicating that the arteriosclerosis subtypes do not significantly differ in their impact on MCA pulsatility. Research shows that IEL calcification and intimal calcification may contribute to increased MCA pulsatility through different mechanisms. IEL calcification is associated with arterial stiffness [29], which is thought to increase pulsatility through elastic fiber stress and greater load bearing [39]. How intimal calcification may influence pulsatility is less well understood. Previous studies suggest that it may compromise arterial distensibility beyond the site of the calcified plaque, though the underlying mechanisms remain elusive [40]. Due to their shared contribution to vascular rigidity and altered hemodynamics, the overall effect of IEL and intimal calcification on MCA pulsatility may appear similar.

Both ICAC and VAC were consistently associated with MCA pulsatility, but not with blood flow velocity. We initially hypothesized that ICAC and VAC presence and burden would be associated with lower mean blood flow velocity. This hypothesis was partly based on previous research indicating that atherosclerosis in the cerebral vasculature is associated with lower cerebral blood flow in a community-based, younger sample (mean age 56.2 years) [41]. While blood flow velocity (i.e. unidirectional blood displacement per unit of time) and cerebral blood flow (i.e. blood volume per unit of tissue volume per unit of time) are distinct hemodynamic measures, previous studies indicate that they are highly correlated [42, 43]. The absence of an association between arteriosclerosis and downstream blood flow velocity may, therefore, be partly attributed to cerebral autoregulation, defined as the ability to maintain cerebral blood flow irrespective of variations in arterial blood pressure [44]. This interpretation aligns with our correlation analysis, which indicated only a weak association between blood flow velocity in the MCA and basal ganglia and no correlation with the semioval center.

Our study did not find an association between intracranial arteriosclerosis and pulsatility and blood flow velocity in the small perforating arteries further downstream. Previous research shows that dynamic damping of pulsatility between the MCA and the small perforating arteries is enhanced in relation to proximal stress (i.e. higher proximal pressure and arterial stiffness) [45]. Therefore, dynamic pulsatility damping between the MCA and the small perforating arteries may explain the absence of a direct association between intracranial arteriosclerosis and small vessel pulsatility. Another possible explanation is the relatively healthy profile of our study population. As noted in the sensitivity analysis, the prevalence of hypertension and intracranial arteriosclerosis was markedly lower in our sample compared to the age- and se-matched sample from the Rotterdam Study (Table S2). This suggests that ICAC and VAC may only affect small vessel function in individuals with more advanced disease, not in healthier aging individuals.

Our analyses revealed no statistically significant associations between ICAC or VAC and whole-brain CVR. Nonetheless, it is worth noting that point estimates for cortical and subcortical gray matter as well as white matter hyperintensities indicated a potential negative association. This trend is also reflected in the unadjusted correlation coefficients. Large vessel arteriosclerosis may influence downstream cerebrovascular reactivity through reduced vessel elasticity or impaired endothelial function [46]. These changes could compromise the ability of small perforating arteries to respond adequately to metabolic demands, potentially explaining the observed trend. The lack of statistical significance does not rule out a relationship between intracranial arteriosclerosis and CVR, as the trends might indicate a subtle effect possibly requiring a larger sample size. However, some studies show that CVR can be maintained despite atherosclerotic disease [47] and arterial stiffness [48]. Hence, further research is necessary to elucidate the true nature of this relationship.

The strengths of our study include the use of 7T MRI, which allowed for detailed investigation of different complementary aspects of vessel function both in the MCA and the microvasculature. Compared to previous 7T studies [49, 50], our sample size is relatively large and the strong correlations between measures of the same modality further indicate that our 7T MRI measures are internally consistent with a good signal-to-noise ratio. Additionally, the homogeneity of our study sample enhances the internal validity of our findings.

While the homogeneous nature of our sample is a strength, it also limits the generalizability of our results to a broader, more diverse population, particularly those with poorer health. As indicated by our sensitivity analysis, prevalence of both arteriosclerosis and cardiovascular risk factors was lower in this sample compared to previous Rotterdam Study cohorts. Therefore, it is possible that our study was subject to selection bias.

Additionally, there was a time gap of approximately two years between the CT assessment and the 7T MRI scan. Although previous studies suggest that ICAC volumes remain relatively stable over this time period [51], ICAC prevalence most likely increases. As a result, our study may have underestimated the full impact of intracranial arteriosclerosis on MCA and small vessel function.

Another limitation is the cross-sectional study design, which does not allow for drawing conclusions regarding potential longitudinal effects of intracranial arteriosclerosis on small vessel function. Hence, longitudinal studies are needed to better understand the underlying temporal dynamics.

## Conclusion

Our findings suggest that intracranial arteriosclerosis only influences pulsatility in adjacent arteries, but not further downstream in the small perforating arteries. Moreover, we found no association between intracranial arteriosclerosis and blood flow velocity. Although there may be a subtle effect of intracranial arteriosclerosis on whole-brain CVR, this relationship remains uncertain. Taken together, the findings underscore the need for further research to identify the pathways underlying intracranial arteriosclerosis and cerebrovascular function.

## Data Availability

Data can be obtained upon request. Requests should be directed towards the management team of the Rotterdam Study, which has a protocol for approving data requests. Because of restrictions based on privacy regulations and informed consent of the participants, data cannot be made freely available in a public repository.

## ACKNOWLEDGMENTES

We would like to acknowledge the immense contribution of the data management team of the Rotterdam Study, with Jolande Verkroost-van Heemst in particular, and of the Imaging Trialbureau of the department of Radiology and Nuclear Medicine. We acknowledge Danique Versluis, Philippe-Jan Ramondt, Charlotte Verhagen, Guido Camma, Alessandro Nigi, Keira Vriend, and Madouc Linders for their efforts in acquisition of all the data. We also want to acknowledge Madouc and Beatrijs Grotenhuis for their help in the data analysis.

