## Supplement for "Intracranial arteriosclerosis and cerebrovascular function in the general aging population – A 7T MRI Study"

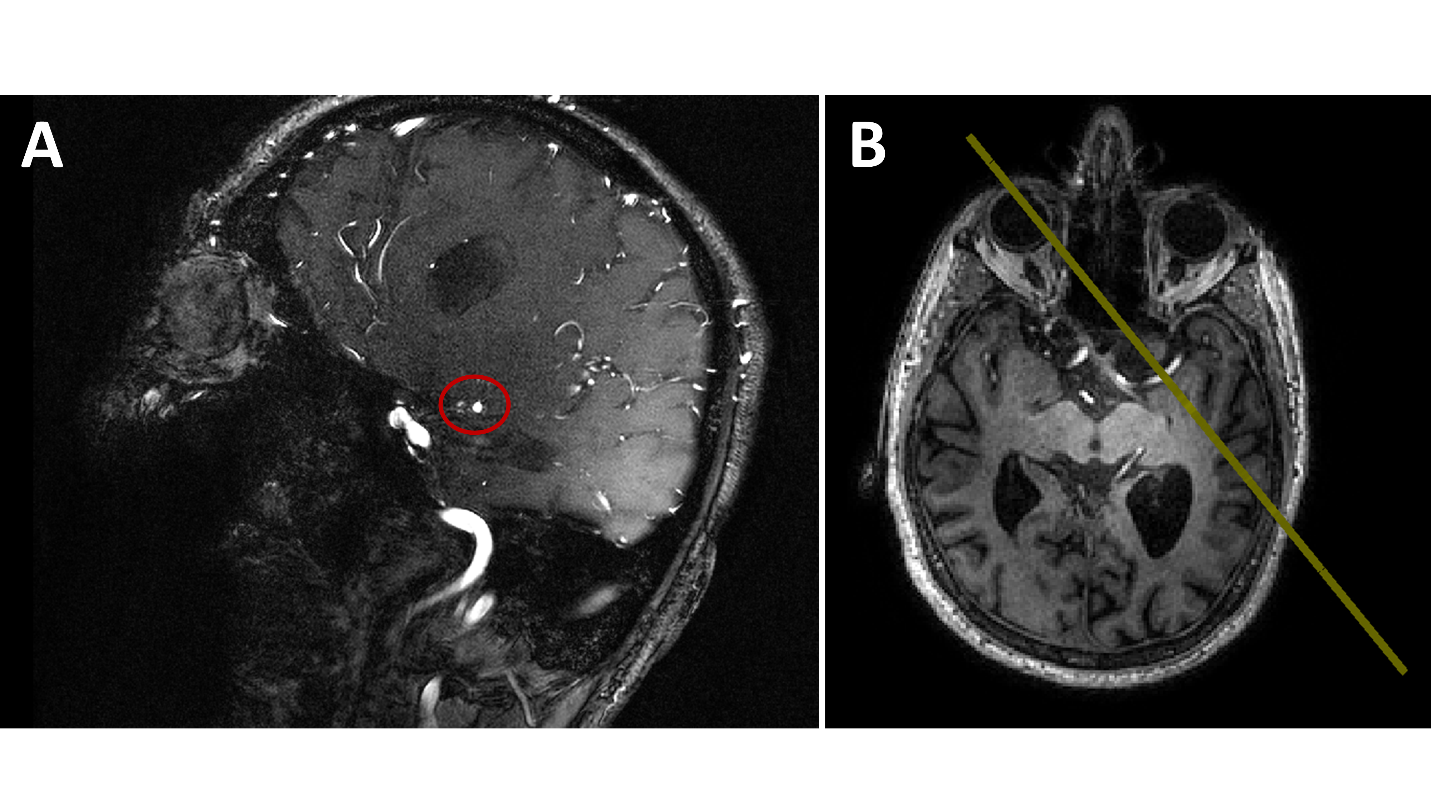
**Figure S1.** (A) Sagittal view on the brain with the middle cerebral artery (circled in red) located in the center. (B) Single 2D slice (yellow) planned perpendicularly on the middle cerebral artery for region of interest delineation.

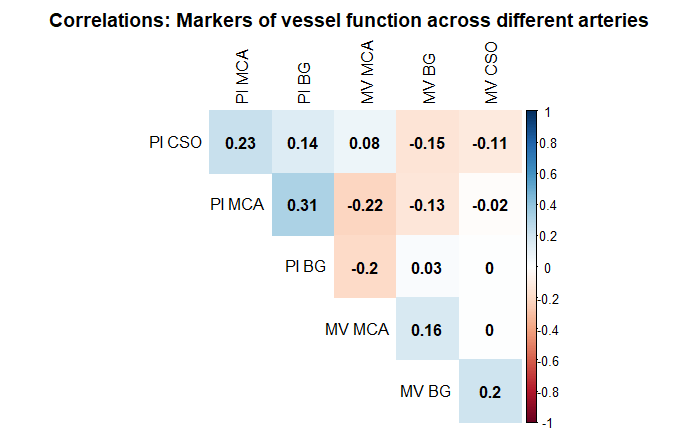

**Figure S2.** Unadjusted correlation coefficients for pulsatility (PI) and mean-blood flow velocity (MV) in the middle cerebral artery (MCA), basal ganglia (BG), and semioval center (CSO).

**Table S1.** Linear regression results investigating the relationship between intracranial arteriosclerosis and middle cerebral artery pulsatility and blood flow velocity.

|  | Pulsatility | | Blood flow velocity | |
| --- | --- | --- | --- | --- |
| Coefficient | Beta | 95% CI | Beta | 95% CI |
| ICAC presence | 0.08 | 0.02 - 0.13 | 1.09 | -2.58 - 4.75 |
| ICAC tertiles |  |  |  |  |
| 1 | 0.05 | -0.02 - 0.11 | 1.36 | -2.95 – 5.67 |
| 2 | 0.08 | 0.02 - 0.14 | -0.58 | -4.84 - 3.67 |
| 3 | 0.11 | 0.05 - 0.18 | 3.57 | -1.16 - 8.29 |
| ICAC subtypes |  |  |  |  |
| Intimal | 0.06 | 0.00 – 0.12 | 1.29 | -2.59 – 5.17 |
| IEL | 0.11 | 0.03 – 0.18 | -0.97 | -5.93 – 3.98 |
| Mixed | 0.10 | 0.02 – 0.17 | 2.86 | -2.39 – 8.12 |
| VAC presence | 0.08 | 0.00 - 0.15 | -1.64 | -6.75 – 3.46 |
| VAC volume | 0.00 | 0.00 - 0.00 | -0.04 | -0.15 - 0.07 |
| *Note.* ICAC=intracranial carotid artery calcification; VAC=vertebral artery calcification; IEL=internal elastic lamina calcification. Betas indicate the change in mean blood flow velocity (cm/s) for categorical calcification estimates compared to no calcification or per 1mm^3^ increase of calcification. All models have been adjusted for age and sex. | | | | |

**Table S2.** Linear mixed model results showing the relationship between intracranial arteriosclerosis and pulsatility in the basal ganglia and semioval center.

|  | Basal ganglia | | Semioval center | |
| --- | --- | --- | --- | --- |
| Coefficient | Beta | 95% CI | Beta | 95% CI |
| ICAC presence | 0.02 | -0.02 – 0.06 | -0.00 | -0.04 – 0.04 |
| ICAC tertiles |  |  |  |  |
| 1 | 0.02 | -0.03 – 0.06 | -0.01 | -0.06 – 0.04 |
| 2 | 0.03 | -0.02 – 0.08 | 0.01 | -0.04 – 0.06 |
| 3 | -0.00 | -0.06 – 0.05 | -0.01 | -0.07 – 0.05 |
| ICAC subtypes |  |  |  |  |
| Intimal | 0.01 | -0.03 – 0.05 | 0.01 | -0.04 – 0.05 |
| IEL | 0.03 | -0.02 – 0.08 | -0.03 | -0.09 – 0.02 |
| VAC presence | -0.01 | -0.08 – 0.06 | 0.05 | -0.02 – 0.13 |
| VAC volume | 0.00 | -0.00 – 0.00 | 0.00 | -0.00 – 0.00 |
| Observations | 366 | | 348 | |
| Participants | 183 | | 174 | |

*Note.* ICAC=intracranial carotid artery calcification; IEL=internal elastic lamina calcification; VAC=vertebral artery calcification. Betas indicate the change in pulsatility ((V_max_-V_min_)/V_mean_) for categorical calcification estimates compared to no calcification or per 1mm^3^ increase of calcification. All models have been adjusted for age and sex.

**Table S3.** Linear mixed model results showing the relationship between intracranial arteriosclerosis and blood flow velocity in the basal ganglia and semioval center.

|  | Basal ganglia | | Semioval center | |
| --- | --- | --- | --- | --- |
| Coefficient | Beta | 95% CI | Beta | 95% CI |
| ICAC presence | 0.01 | -0.21 – 0.22 | 0.03 | -0.01 – 0.08 |
| ICAC tertiles |  |  |  |  |
| 1 | 0.05 | -0.20 – 0.30 | 0.02 | -0.03 – 0.08 |
| 2 | 0.11 | -0.15 – 0.37 | 0.05 | -0.00 – 0.11 |
| 3 | -0.24 | -0.53 – 0.05 | 0.03 | -0.04 – 0.10 |
| ICAC subtypes |  |  |  |  |
| Intimal | 0.06 | -0.16 – 0.28 | 0.02 | -0.02 – 0.07 |
| IEL | -0.10 | -0.38 – 0.18 | 0.05 | -0.01 – 0.11 |
| VAC presence | -0.12 | -0.49 – 0.26 | 0.08 | 0.00 – 0.16 |
| VAC volume | -0.01 | -0.02 – 0.00 | 0.00 | -0.00 – 0.00 |
| Observations | 366 | | 348 | |
| Participants | 183 | | 174 | |

*Note.* ICAC=intracranial carotid artery calcification; VAC=vertebral artery calcification; IEL=internal elastic lamina calcification. Betas indicate the change in mean blood flow velocity (cm/s) for categorical calcification estimates compared to no calcification or per 1mm^3^ increase of calcification. All models have been adjusted for age and sex.

**Table S4.** Linear mixed model results showing the relationship between intracranial arteriosclerosis and cerebrovascular reactivity.

|  | Cortical gray matter | | Subcortical gray matter | | White matter | | Normal-appearing white matter | | White matter hyperintensities | |
| --- | --- | --- | --- | --- | --- | --- | --- | --- | --- | --- |
| Coefficient | Beta | 95% CI | Beta | 95% CI | Beta | 95% CI | Beta | 95% CI | Beta | 95% CI |
| ICAC presence | -0.003 | -0.014 – 0.007 | -0.008 | -0.022 – 0.005 | 0.001 | -0.007 – 0.010 | 0.001 | -0.007 – 0.010 | -0.023 | -0.064 – 0.017 |
| ICAC tertiles |  |  |  |  |  |  |  |  |  |  |
| 1 | -0.001 | -0.012 – 0.010 | -0.006 | -0.020 – 0.008 | 0.003 | -0.006 – 0.012 | 0.003 | -0.006 – 0.012 | -0.022 | -0.066 – 0.022 |
| 2 | -0.008 | -0.020 – 0.004 | -0.014 | -0.030 – 0.002 | -0.002 | -0.012 – 0.008 | -0.003 | -0.013 – 0.007 | -0.026 | -0.076 – 0.024 |
| 3 | -0.009 | -0.025 – 0.008 | -0.002 | -0.023 – 0.020 | -0.000 | -0.013 – 0.012 | -0.000 | -0.013 – 0.012 | -0.025 | -0.083 – 0.032 |
| ICAC subtypes |  |  |  |  |  |  |  |  |  |  |
| Intimal | -0.003 | -0.014 – 0.008 | -0.009 | -0.023 – 0.005 | 0.000 | -0.008 – 0.009 | 0.000 | -0.008 – 0.009 | -0.025 | -0.067 – 0.018 |
| IEL | -0.001 | -0.015 – 0.012 | -0.004 | -0.022 – 0.013 | 0.005 | -0.006 – 0.015 | 0.005 | -0.006 – 0.015 | -0.020 | -0.074 – 0.034 |
| VAC presence | -0.006 | -0.022 – 0.009 | -0.002 | -0.022 – 0.018 | 0.004 | -0.009 – 0.016 | 0.004 | -0.008 – 0.017 | -0.023 | -0.092 – 0.046 |
| VAC volume | -0.000 | -0.001 – 0.000 | 0.000 | -0.000 – 0.001 | 0.000 | -0.000 – 0.000 | 0.000 | -0.000 – 0.001 | -0.035 | -0.083 – 0.013 |
| Observations | 340 | | 340 | | 321 |  | 321 |  | 219 |  |
| Participants | 170 | | 170 | | 164 |  | 164 |  | 143 |  |

Note. ICAC=intracranial carotid artery calcification; VAC=vertebral artery calcification; IEL=internal elastic lamina calcification. Betas indicate the change in cerebrovascular reactivity for categorical calcification estimates compared to no calcification or per 1mm3 increase of calcification. All models have been adjusted for age and sex.

**Table S5.** Comparison of clinical characteristics of the current sample (7T MRI sample) with an age- and sex-matched sample from the Rotterdam study (RS CT sample)

| Characteristics | 7T MRI (N=195) | RS CT (N=195) |
| --- | --- | --- |
| Age at CT (mean, SD) | 68.6 (4.69) | 67.8 (7.10) |
| Sex (n female, %) | 85 (43.6%) | 85 (43.6%) |
| Body mass index (mean, SD)  Missing (n, %) | 27.4 (4.1)  0 (0%) | 28.1 (3.8)  1 (0.5%) |
| Diabetes (n, %) | 34 (17.4%) | 31 (15.9%) |
| Smoking (n, %)  Current  Former  Never  Missing (n, %) | 15 (7.1%)  121 (62.1%)  59 (30.3%)  1 (0.5%) | 40 (20.5%)  102 (52.3%)  52 (26.7%)  0 (0%) |
| Hypertension (n, %) | 88 (45.1%) | 143 (73.3%) |
| Dyslipidemia (n, %)  Missing (n, %) | 37 (19.0%)  0 (0%) | 47 (24.1%)  2 (1.0%) |
| ICAC  Prevalence (n, %)  Volume [median, IQR] | 151 (77.4%)  48.8 [168] | 162 (83.1%)  41.1 [98.6] |
| VAC  Prevalence (n, %)  Volume [min, max] | 20 (10.3%)  [0, 129] | 38 (19.5%)  [0, 88] |

*Note.* RS=Rotterdam Study; ICAC=intracranial artery calcification; VAC=vertebral artery calcification
